# Investigating CAR-T Treatment Access for Multiple Myeloma Patients Using Real-World Evidence

**DOI:** 10.1101/2025.04.16.25325784

**Authors:** Jaysón Davidson, Anupama Kumar, Ayan Patel, Irene Y. Chen, Atul J. Butte, Travis Zack

## Abstract

**Objective:** Multiple myeloma (MM) is the second most common hematologic malignancy in the U.S., with Black patients being diagnosed at twice the rate of White patients. MM treatment options are limited and ineffective, but CAR-T therapies show promise. However, their limited availability results in disparities in access. This study aimed to explore disparities in Multiple Myeloma disease risk and CAR-T therapy access.

**Methods:** Our study included a population-based cohort of 12,360 patients diagnosed with Multiple Myeloma who received more than one cancer therapy extracted from the University of California Health Data Warehouse (UCHDW) between January 2021 and November 2024. Regression models were used to compute odds ratio (OR) and 95% confidence intervals (CI) associating disease severity, UC- Location, and baseline demographics with CAR-T therapy access. The GPT-4 inference model was prompted with a zero-shot learning approach to analyze UCSF clinical notes with the following objectives: (1) Was CAR-T discussed? [yes/no], (2) Is the patient eligible for CAR-T? [yes/no/unclear], and (3) Provide the rationale for the eligibility determination.

**Results:** Our study included 12,360 patients (mean age 68.5 years, SD 12.8 years) treated for multiple myeloma across the University of California Health System, 320 of which received CAR-T (Table-1). Overall, 51.6% of MM patients identified as Male, and 48.4% as Female. Disease Severity was measured by the International Staging System (ISS) and was distributed by ISS Stage: I (65.3%), II (24.4%), III (2.8%), and None (7.5%). Patients treated at UC-1 (49.3%), and UC-2 (50.0%) were primarily diagnosed with Stage II, while patients at UC-3 (55.5%) were primarily diagnosed with Stage I. Our model indicated that patients who identified as Black or African American (OR= 0.33, [95% CI, 0.17- 0.62) were less likely to receive CAR-T therapy when compared to White patients. Patients treated at UC-3 with predominantly Black or African American patients (OR = 0.42, [95% CI, 0.30-0.59]) were less likely to receive CAR-T therapy when compared to UC-1. We identified CAR-T eligibility for 270 UCSF patients and found those who identified as other Pacific Islander had the highest rate of eligibility without discussions at 50%, followed by Black or African American (4.2%), Asian (3.2%), and White (0.6%).

**Conclusion and Relevance:** This study emphasizes the influence of race and UC-Location on disparities in CAR-T therapy access.

## Introduction

Multiple myeloma (MM) is a hematologic malignancy characterized by the clonal proliferation of malignant plasma cells, which can result in end-organ damage, morbidity, and mortality (with a five-year overall survival of 58%)^1^. Black patients are diagnosed at twice the rate and, on average, 5-10 years earlier than White patients^2^ and are more likely to present with anemia, hypercalcemia, and kidney dysfunction^3,4^. They are underrepresented in clinical trials investigating novel therapies^5,6^; despite recent real-world studies showing they achieve better survival rates than White patients when given equal access to care^6–9^. Hispanic or Latino patients with myeloma similarly have a higher incidence and younger age of diagnosis. These patients are known to have longer time intervals from diagnosis to therapy initiation and lower utilization of essential therapies such as autologous stem cell transplant (ASCT)^10^.

Standard of care front-line therapy consists of triplet or quadruplet induction therapy, followed by ASCT consolidation and maintenance in eligible patients; transplant-ineligible patients receive attenuated ongoing systemic therapy. ^11–16^ Despite advances, multiple myeloma invariably relapses. A recent breakthrough in the treatment of relapsed/refractory multiple myeloma (RRMM) is the use of chimeric antigen receptor T-cell (CAR-T) therapy, which are engineered T-cells designed to target B-cell maturation antigen (BCMA) on plasma cells^17,18^. Two FDA-approved BCMA CAR-T therapies in MM, idecabtagene vicleucel (idecel) and ciltacabtagene autoleucel (ciltacel), are now approved for both early and late relapse. These therapies have demonstrated unprecedented outcomes, including a 98% overall response rate (ORR) and nearly 36-month progression-free survival (PFS) with ciltacel^19^, and offer the appeal of a treatment-free interval (TFI), but they require access to a limited number of academic centers with expertise in CAR-T management and related toxicities. Patients lacking access to these centers, the ability to relocate, or adequate social support for a caregiver may face barriers to receiving CAR-T therapy. While these barriers may disproportionately affect Black, and Hispanic or Latino patients, the extent to which they influence CAR-T therapy access remains poorly understood.

Therefore, we hypothesize that a patient’s location and race are strongly associated with CAR-T therapy access for MM patients, considering both disease risk and baseline demographics. To test this hypothesis, our study systematically analyzed the association between access to CAR-T and disease risk, Social Determinants of Health (SDOH), and Race for MM patients. To do so, we leveraged a clinical data warehouse containing over 9 million individuals across the three main academic medical centers that offer CAR-T administration for cancer patients within the University of California (UC) Health system: UCLA, UCSD, and UCSF.

## Methods

### Data Supporting Study: University of California Health Data Warehouse (UCHDW)

Data for this study was drawn from the University of California Health System, which includes 20 health professional schools (6 medical schools), 5 academic health centers (UC San Francisco, UC Los Angeles, UC Davis, UC Irvine, and UC San Diego), and 12 hospitals. It has built a secure central data warehouse (UCHDW) for operational improvement, promotion of quality patient care, enabling the next generation of clinical research^20^. The repository currently holds data securely on over 9 million patients seen since 2012. EHR data is extracted from each site and transformed into vendor neutral Observational Medical Outcomes Partnership (OMOP) common data model^21^. De-identification of the data has already been completed to enable clinical research projects, under guidance from UC campus institutional review boards, privacy, and compliance. Research use of UCHDW is considered non-human subject’s research.

### Study Population

UCHDW data was extracted from patients diagnosed with MM ICD-10 code (C90.0) and had documentation for multiple cancer therapies and procedures from EHRs spanning from the year 2012 up to August 2024 **(Supplement-1).** To select a cohort of patients with potential CAR-T therapy, we used the following inclusion/exclusion criteria to create a final study cohort of 12,360 patients (Figure 1). If patients diagnosed with MM did not have primary insurance information, they were excluded from our study to include only patients with insurance. We also excluded patients who didn’t have and those who didn’t have an Area Deprivation Index (ADI) value. We further excluded patients without a visit on or after January 1, 2021. Because eligibility for non-investigational CAR-T currently requires at least one previous line of therapy, patients who did not have more than one cancer treatment were excluded from our study. We considered all therapies patients received after MM diagnosis, with at least one year of treatment. Patients who were treated at a UC-facility after January 2021 that offered CAR-T therapies were included in our study. Finally, all patients under 18 years of age at the time of diagnosis were excluded from the cohort. This study followed the Strengthening the Reporting of Observational Studies in Epidemiology (STROBE) reporting guidelines to ensure the quality of our observational study^22^.

**Figure 1:**
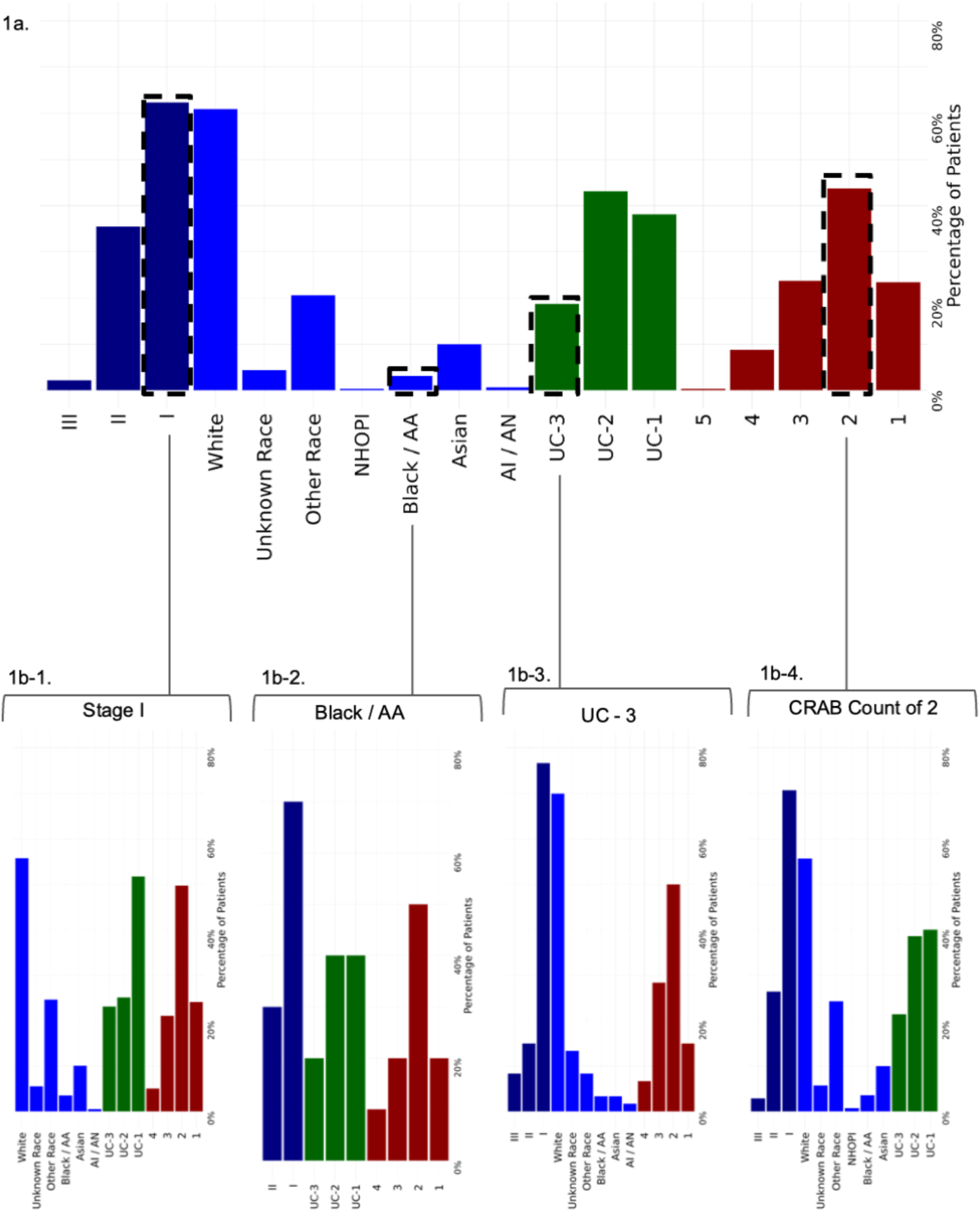
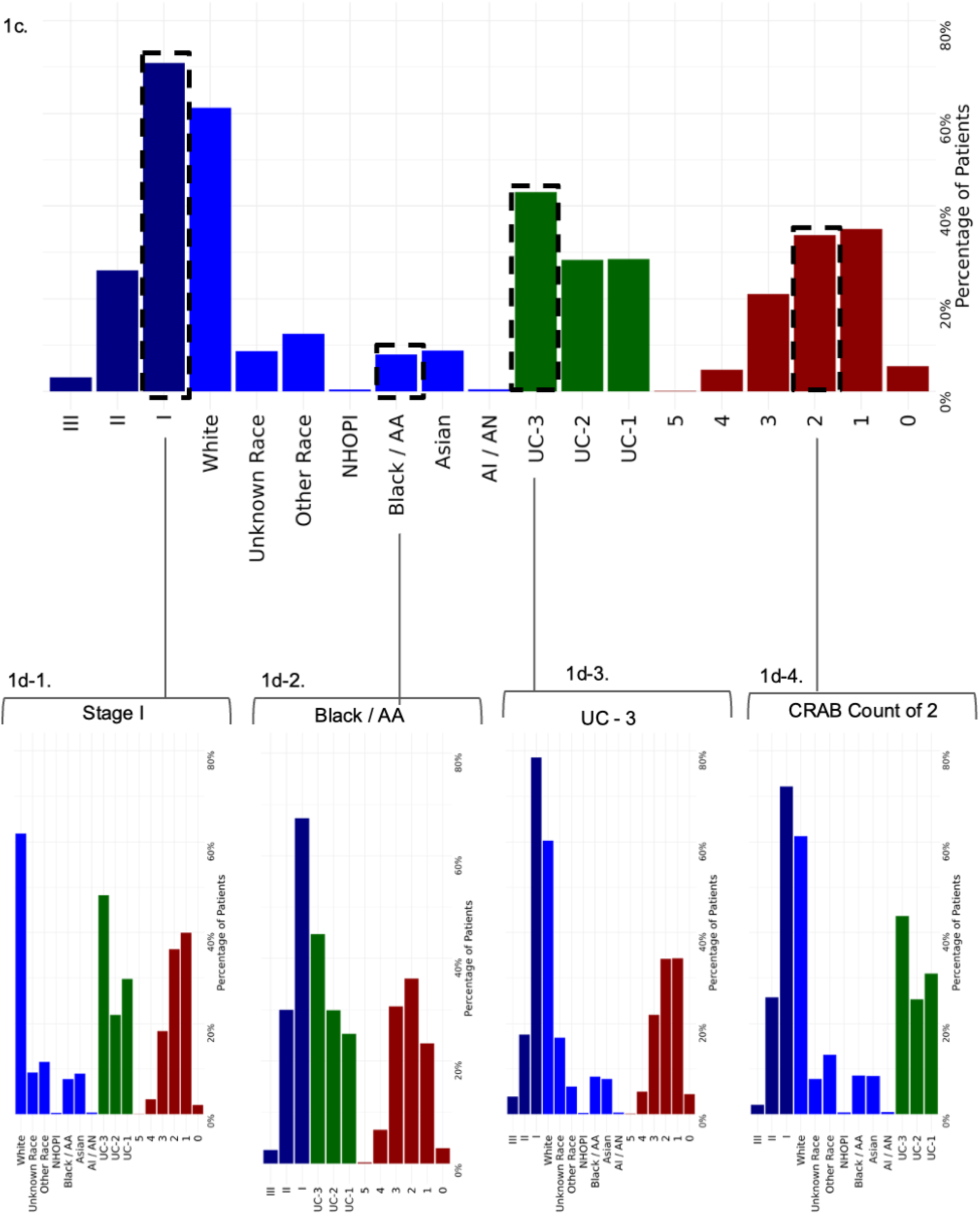
1a. Distribution of demographics for patients who received CAR-T therapy. 1b-1. Patients diagnosed with ISS Stage I. 1b-2. Patients who identified as Black or African American. 1b-3. Patients who were treated at UC-3. 1b-4 Patients with 2 CRAB features. 1c. Distribution of demographics for patients who did not receive CAR-T therapy. 1d-1. Patients diagnosed with ISS Stage I. 1d-2. Patients who identified as Black or African American. 1d-3. Patients who were treated at UC-3. 1d-4 Patients with 2 CRAB features

### Area Deprivation Index (ADI)

To estimate patient-level SDOH, we employed ADI as a proxy for patients socioeconomic status^23^. ADI does not account for specific individual or family characteristics, but it is a general estimate of SDOH based on home location census tract characteristics by integrating data from 17 different variables sourced from the U.S. Census, with factors related to poverty, housing, employment, and education. ADI as a metric has been used in previous observational studies^24–28^ to understand its SDOH role on health outcomes in other disease states, distinguishing the most deprived areas with higher percentile scores from the least deprived areas with lower percentile scores^23,29–31^.

### Study Variables

Patients were classified as receiving CAR-T or not through data collected by UC Health as above. Additional variables included ADI, Age (in years), patient-reported Sex (Male, Female), presence of primary insurance coverage, patient-reported Race, Ethnicity, UC-Location, and International Staging System (ISS) Disease Stage. Number of (CRAB) features were identified for patients with C: Hypercalcemia, R: Renal failure, A: Anemia, or B: Bone disease, or bone pain. To identify cancer procedures and therapies for patients, we categorized all treatments (surgery, chemotherapy, radiation, ASCT, monoclonal antibodies, corticosteroids, alkylators, bispecific antibodies, CAR-T, immunomodulatory agents, nuclear export inhibitors, proteasome inhibitors to ensure that we correctly accounted for cancer therapeutic classes before CAR-T treatments. The International Staging System (ISS) for MM disease severity was calculated for patients by measuring the patient’s serum albumin, and serum β2 microglobulin (Sβ2M) levels^32^. Patients were categorized as ISS Stage I, II, III, or Unknown. Age (in years) was calculated relative to the index date from the patients’ date of birth. Patients with primary insurance healthcare coverage were utilized as Medicare, Medicaid, Veterans Affairs, or private insurance types. Patient reported race was captured as White, Asian, Black or African American, Native Hawaiian or Pacific Islander, American Indian or Alaska Native, Multi Race, Other Race, or Unknown. Patients reported ethnicity was utilized as Hispanic or Latino, Not Hispanic -or Latino or Unknown. UC-Location was identified as patients treated at UC-1, UC-2, UC-3.

### Statistical Analysis

We conducted our data analysis from March 2024 to December 2024. Descriptive statistics were calculated first for each covariate and secondly, for the overall patient cohort, by using Analysis of Variance (ANOVA), and chi-squared tests^33,34^. We employed a Generalized Linear Model (GLM) to estimate the association of primary insurance coverage, race, and ethnicity and our covariates on our key outcome: CAR-T therapy access^33,34^. We adjusted for age (in years), sex (male, female), ADI, Primary Insurance coverage, UC-Location, number of CRAB features, ISS Disease Stage, race, and ethnicity. Association and its strength were reported as odds ratio (OR) with 95% confidence Intervals (CI) including p-values indicating the statistical significance of how likely a patient was to receive CAR-T therapy. An OR was considered significant if its 95% CI did not span 1, and p-value < 0.05. An OR = 1 indicates no difference between receiving CAR-T or not, OR > 1 indicates increased likelihood of a patient receiving CAR-T, and OR < 1 indicates decreased likelihood of a patient receiving CAR-T. All the calculations were performed using R statistical software version 3.6.3 (R Project for Statistical Computing).

### CAR-T Eligibility Identification through Large Language Models

To identify CAR-T eligibility for patients from the UCSF de-identified clinical database, we applied the inclusion and exclusion criteria outlined in the study population section of the methods. We excluded patients who did not have clinical notes associated with their diagnoses. Additionally, patients whose clinical notes had an index date on or before January 1, 2021, were excluded. We also excluded patients with clinical notes that were not within 365 days of the index date of their last cancer therapy or procedure, resulting in a final cohort of 270 patients with 417 clinical notes (Supplement-3). To identify CAR-T eligibility, we utilized the GPT-4 inference model with a zero-shot learning approach. GPT-4 was prompted to analyze the “Assessment & Plan” section of the clinical notes written by physicians. The model was tasked with four objectives: (1) Was CAR-T discussed? [yes/no], (2) Is the patient eligible for CAR-T? [yes/no/unclear], (3) Provide the rationale for the eligibility determination, and (4) Classify the eligibility rationale for CAR-T discussions into predefined groups: (“Ineligible due to disease criteria”, “Ineligible due to frailty or comorbidities”, “Ineligible due to social determinants of health”, “Ineligible due to not enough prior lines of therapy”, “Eligible now or potentially eligible in future”, “Not enough information to determine eligibility”, and “Other”). The model’s accuracy was tested and validated on a subsample of clinical notes by comparing its outputs against the ground truth to assess its performance in identifying CAR-T eligibility. GPT-4 model outputs were analyzed and checked for accuracy by clinicians. The identified CAR-T eligibility outputs were then visualized and presented in a tabular format.

## Results

Our study included 12,360 patients (mean age 68.5 years, SD 12.8 years) treated for multiple myeloma across the University of California, 320 of which received CAR-T (Table 1). Of these patients, 51.6% of MM patients identified as Male, and 48.4% as Female. 61.2% of patients identified as White, while 7.8% identified as Black or African American and 14.9% as Hispanic or Latino. Among patients receiving CAR-T, 57.5% were Male and 60.9% identified as White. Patients treated at each UC-Location were primarily White, but UC-3 had a higher distribution of patients who identified as Black, and Unknown, while UC-1 had a higher distribution of patients who identified as Unknown Race, and Hispanic or Latino.

**Table 1:** Baseline Characteristics of Multiple Myeloma Patients.

| Did a patient receive CAR-T therapy? |  |  |  |
| --- | --- | --- | --- |
| Variable | Didn't receive CAR-T therapy | Received CAR-T therapy | Total |
|  | (N=12040) | (N=320) | (N=12360) |
| Age |  |  |  |
| Mean (SD) | 68.7 (12.7) | 63.1 (12.0) | 68.5 (12.8) |
| Area Deprivation Index (ADI) |  |  |  |
| Mean (SD) | 3.88 (2.64) | 4.28 (2.82) | 3.89 (2.65) |
| Number of CRAB Features |  |  |  |
| Mean (SD) | 1.85 (0.976) | 2.19 (0.904) | 1.86 (0.975) |
| Sex |  |  |  |
| Female | 5852 (48.6%) | 136 (42.5%) | 5988 (48.4%) |
| Male | 6188 (51.4%) | 184 (57.5%) | 6372 (51.6%) |
| Race |  |  |  |
| American Indian or Alaska Native | 51 (0.4%) | <10 (<10%) | 53 (0.4%) |
| Asian | 1062 (8.8%) | 32 (10.0%) | 1094 (8.9%) |
| Black or African American | 959 (8.0%) | 10 (3.1%) | 969 (7.8%) |
| Native Hawaiian or Other Pacific Islander | 46 (0.4%) | <10 (<10%) | 47 (0.4%) |
| Other Race | 1501 (12.5%) | 66 (20.6%) | 1567 (12.7%) |
| Unknown | 1046 (8.7%) | 14 (4.4%) | 1060 (8.6%) |
| White | 7375 (61.3%) | 195 (60.9%) | 7570 (61.2%) |
| Ethnicity |  |  |  |
| Hispanic or Latino | 1768 (14.7%) | 78 (24.4%) | 1846 (14.9%) |
| Not Hispanic or Latino | 9305 (77.3%) | 236 (73.8%) | 9541 (77.2%) |
| Unknown | 967 (8.0%) | <10 (<10%) | 973 (7.9%) |
| ISS Stage |  |  |  |
| I | 7878 (65.4%) | 199 (62.2%) | 8077 (65.3%) |
| II | 2903 (24.1%) | 113 (35.3%) | 3016 (24.4%) |
| III | 336 (2.8%) | <10 (<10%) | 343 (2.8%) |
| None | 923 (7.7%) | <10 (<10%) | 924 (7.5%) |
| UC-Location |  |  |  |
| UC-1 | 3442 (28.6%) | 122 (38.1%) | 3564 (28.8%) |
| UC-2 | 3419 (28.4%) | 138 (43.1%) | 3557 (28.8%) |
| UC-3 | 5179 (43.0%) | 60 (18.8%) | 5239 (42.4%) |
| Primary Insurance Coverage |  |  |  |
| Medicaid | 1611 (13.4%) | 53 (16.6%) | 1664 (13.5%) |
| Medicare | 4775 (39.7%) | 106 (33.1%) | 4881 (39.5%) |
| Private | 5394 (44.8%) | 156 (48.8%) | 5550 (44.9%) |
| VA Insurance | 260 (2.2%) | <10 (<10%) | 265 (2.1%) |

Baseline Characteristics of Multiple Myeloma patients with 1 or more MM therapies between January
2012 - May 2024.

Primary insurance coverage was distributed as Medicaid (13.5%), Medicare (39.5%), Private (44.9%), and Veterans Affairs Insurance (2.1%). Disease Severity was measured by the International Staging System (ISS), with patients distributed by ISS Stage: I (65.3%), II (24.4%), III (2.8%), and None (7.5%). Patients treated at each UC-Location were primarily diagnosed with Stage I but patients at UC-1 (21.4%), and UC-2 (39.2%) had a higher distribution of patients diagnosed with Stage II. We used the number of CRAB features per patient to assess disease severity, the Mean (SD) was 1.86 (0.98), with the most common feature being Anemia **(Supplement-2).**

Using a univariate analysis, we found race to be a significant determinant of CAR-T treatment access (Methods, p < 0.001, χ2 test) (Table 1). ISS Stage was significantly different across Race and was a significant determinant of CAR-T treatment access (Methods, p < 0.001, χ2 test). We found patient’s Socioeconomic status and location to be a significant determinant of CAR-T access (Methods, p = 0.007, ANOVA). Additionally, we modeled the relationship between UC-Location, Race, and CAR-T access and found patients’ UC-Location to be a strong determinant (Methods, p < 0.001, χ2 test). Furthermore, we ran a regression analysis modeling the relationship between receiving CAR-T treatment that adjusted for ISS Stage, UC-Location, race, ethnicity, number of CRAB features, ADI, and primary insurance coverage. The results were highly significant in showing that these variables played a role in CAR-T treatment access for patients (Table 2).

**Table 2:** Results of Logistic Regression Model.

| Variable | Odds Ratio (95%CI) | P-Value |
| --- | --- | --- |
| **Area Deprivation Index (ADI)** | 1.02 (0.97-1.07) | 0.395 |
| **Age** | 0.97 (0.96-0.98) | < 0.001 |
| **UC-Location** |  |  |
| UC-2 | 1.42 (1.09-1.87) | 0.011 |
| UC-3 | 0.42 (0.30-0.59) | < 0.001 |
| **International Staging System (ISS)** |  |  |
| II | 1.15 (0.89-1.48) | 0.271 |
| III | 0.69 (0.32-1.50) | 0.348 |
| None | 0.07 (0.01-0.48) | 0.007 |
| **Number of CRAB Features** | 1.43 (1.27-1.62) | < 0.001 |
| **Race** |  |  |
| American Indian or Alaska Native | 1.04 (0.24-4.43) | 0.959 |
| Asian | 0.99 (0.67-1.46) | 0.959 |
| Black or African American | 0.33 (0.17-0.62) | < 0.001 |
| Native Hawaiian or Other Pacific Islander | 0.77 (0.1-5.72) | 0.802 |
| Other | 1.07 (0.73-1.56) | 0.736 |
| Unknown | 1.26 (0.67-2.37) | 0.47 |
| **Ethnicity** |  |  |
| Hispanic or Latino | 1.26 (0.87-1.81) | 0.216 |
| Unknown | 0.36 (0.15-0.88) | 0.026 |
| **Primary Insurance Coverage** |  |  |
| Medicaid | 0.71 (0.50-1.01) | 0.056 |
| Medicare | 0.96 (0.73-1.25) | 0.752 |
| Veteran Affairs Insurance | 0.61 (0.25-1.53) | 0.292 |

Odds Ratio table of patients receiving CAR-T therapies across patient characteristics. An OR = 1 indicates no difference between receiving or not receiving CAR-T therapies, OR > 1 indicates increased likelihood of receiving a CAR-T therapy and OR < 1 indicates decreased likelihood of receiving CAR-T as a therapy.

Our regression model showed that as patients aged, they were less likely (OR = 0.97, [95% CI, 0.96- 0.98]) to receive CART treatments when compared to younger patients. Patients who identified as Hispanic or Latino (OR = 1.26, [95% CI, 0.87-1.81]) were more likely to receive CAR-T treatment than patients who identified as Not Hispanic or Latino. Most strikingly, patients who identified as Black or African American (OR= 0.33, [95% CI, 0.17-0.62) were highly unlikely to receive CAR-T during MM treatments when compared to White patients. Patients with Private primary insurance coverage were more likely to receive CAR-T over patients with Medicaid (OR = 0.71, [95% CI, 0.50-1.01]), Medicare (OR = 0.96, [95% CI, 0.73-1.25]), and Veteran Affairs (OR = 0.61, [95% CI, 0.25-1.53]) insurance.

Results of our model indicated that disease severity was a strong predictor of CAR-T treatment access for patients. Patients who presented with more CRAB features (OR = 1.43, [95% CI, 1.27-1.62]) were more likely to receive a CAR-T treatment. Patients who were diagnosed with ISS Stage II (OR = 1.15, [95% CI, 0.89-1.48]) were more likely to receive a CAR-T treatment than patients with ISS Stage I MM (Figure 1).

Our results highlighted that UC-Location was a significant determinant of CAR-T treatment administration. Most CAR-T administration for MM patients was done at UC-2 (43.1%). Our model showed that patients treated at UC-3 (OR = 0.42, [95% CI, 0.30-0.59]) were less likely to receive CAR-T therapy when compared to UC-1. We further investigated individual relationships between UC- Location, ISS Stage, and race, by performing additional regression analysis to understand these differences and found significance for race at UC-Locations (p < 0.001), ISS Stage at UC-locations (p < 0.001), and number of CRAB features at UC-Locations (p < 0.001). Interestingly, our results show that CAR-T treatment access is disparate among different population groups based on Race, disease severity, and UC-Location (Figure 2). The distribution of patients within each UC-Location further explained why Black or African American patients, and patients with ISS Stage I were less likely to receive CAR-T (**Supplement-2**). The distribution showed that locations with fewer CAR-T treatments had the highest amount of Black and Hispanic or Latino patients and patients with ISS stage I. This suggests that disparities in access to CAR-T may be driven by access to locations more likely to offer therapy, rather than individual clinical decision-making.

**Figure 2:**
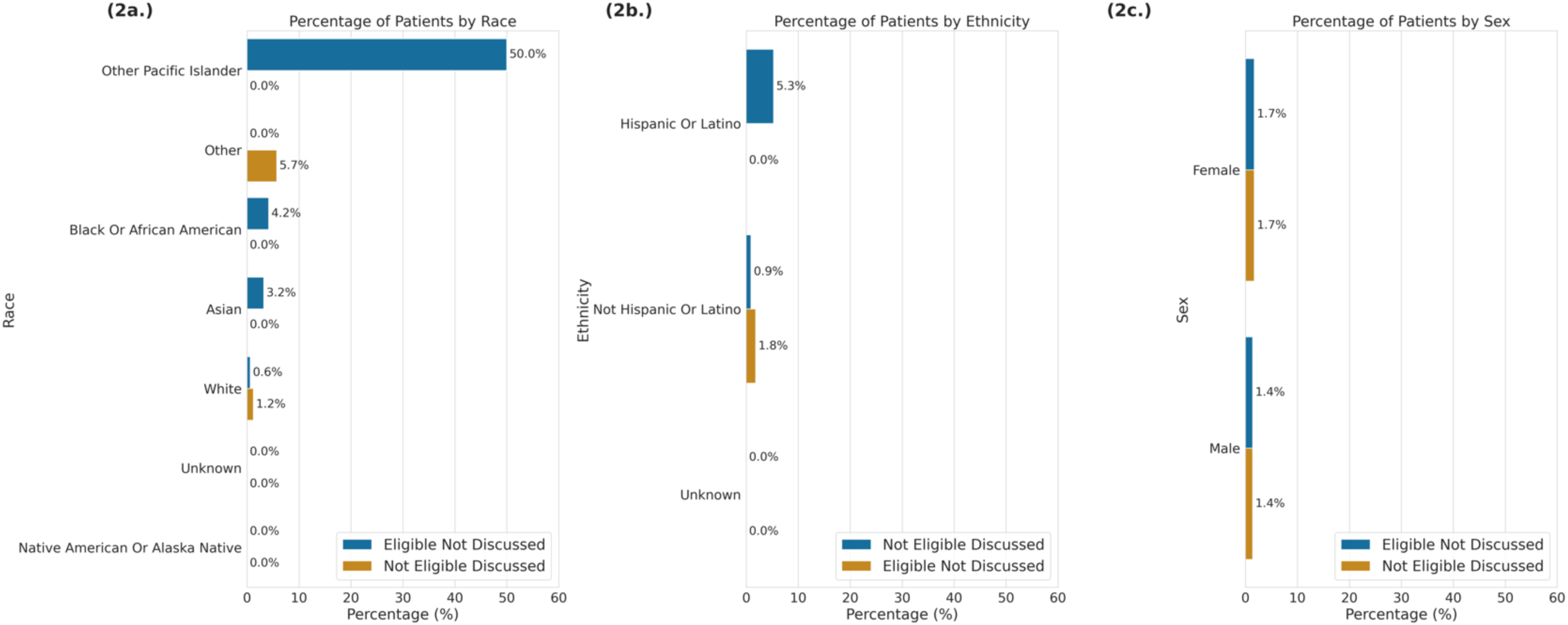
Distribution of UCSF Multiple Myeloma Patients by 2a. Race, 2b. Ethnicity, and 2c. Sex, who were eligible for CAR-T therapy but didn’t have a discussion, and patients who had a discussion but were not eligible for CAR-T therapy.

We further investigated CAR-T therapy access by utilizing a Large Language Model (GPT-4o) deployed within a HIPAA-compliant environment to identify CAR-T eligibility for 270 UCSF patients whose clinical notes contained Assessment & Plan sections. Among these patients, we identified individuals who were considered eligible for CAR-T therapy based on eligibility guidelines but never had a documented discussion about it. We identified CAR-T eligibility for 270 UCSF patients and found patients who identified as other Pacific Islander race had the highest rate of eligibility without discussions at 50%, followed by Black or African American (4.2%), Asian (3.2%), and White (0.6%). These patient populations, despite being eligible for CAR-T therapy, are missing out on potentially life-saving treatments. This finding highlights a critical gap where eligible patients are not being offered or engaged in discussions about CAR-T therapy (Figure 2). For patients who had CAR-T discussions with their provider, we further investigated the topic for patients’ eligibility decisions. Amongst patients with CAR-T discussions we found the LLM classified 33.3% of them to be Eligible now or potentially eligible in the future and 23.3% who were found to be ineligible (Figure 3).

**Figure 3:**
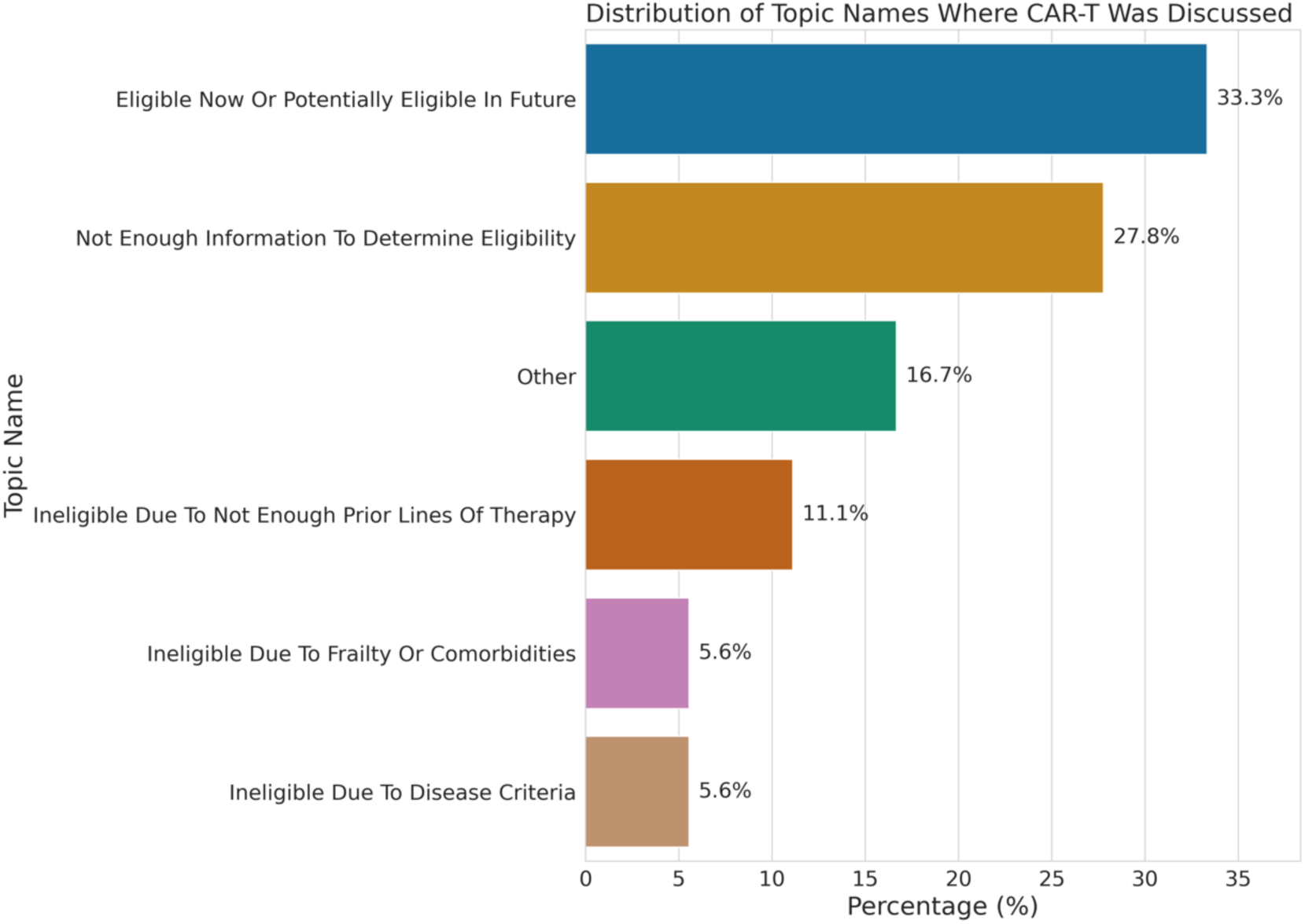
Distribution of topic Modeling for UCSF Multiple Myeloma Patients who had discussions about CAR-T therapy with their provider.

## Discussion

In this retrospective cohort study, we evaluated disease severity, UC-Location, primary insurance coverage, and ADI as independent determinants of CAR-T therapy access for patients diagnosed with MM within the UC-Health system. Notably, we found that therapy access differed by UC-Locations that offered CAR-T administration. Patients at UC-1, and UC-2 which had a higher percentage of White, and Not Hispanic or Latino patients had higher rates of CAR-T therapy administration than patients treated at UC-3. UC-3 has been known to treat patients for both primary and specialty care. Whereas UC-1 and UC-2 are known for specialty care which may be attributed to higher numbers of CAR-T administration. Differences in access by treatment location could also be attributed to the patient’s disease severity.

Our model also revealed disparities in CAR-T access based on patients’ race, ethnicity, and age. First, we found that as patients aged, they were less likely to receive CAR-T. This is aligned with clinical practice, as older patients with RRMM may be preferentially administered bispecific antibodies rather than CAR-T given concerns about risks of high-grade cytokine release syndrome (CRS), immune-effector cell-associated neurotoxicity (ICANS), and delayed neurotoxicity such as Parkinsonism^35,36^. Additionally, we found that patients who identified as Black or African American were unlikely to receive CAR-T therapy compared to White patients. This is significant in showing disparities in CAR-T access, given that Black patients are two times more likely to be diagnosed with MM and have worse survival rates than White patients^14^. We suspect that this may be due to the lack of diversity of patients at UC-1, and UC-2 who have the highest rates of CAR-T administration. This discrepancy may also be related to lower rates of enrollment in CAR-T clinical trials.

Beyond UC-Location and race, we discovered that disease severity played a major role in CAR-T therapy access for patients. Patients who were diagnosed with Stage I, or Stage II diagnosis, were more likely to receive CAR-T therapy than patients with Stage III or ISS Stage None diagnosis. We also found that patients treated at UC-1, and UC-2 had more patients diagnosed with Stage I or Stage II in our dataset. This finding contributes to why UC-1, and UC-2 had more patients with CAR-T therapies than patients at UC-3.

With our understanding of UC-3, primarily treating minorities and patients with lower socioeconomic status, we found it interesting that ADI was not a significant predictor in determining CAR-T access. Considering that socioeconomic status is typically associated with clinical trial access and access to newer treatments in clinical disease, we expected ADI to be significant^38,39^. However, we found it refreshing that Hispanic or Latino patients were more likely to receive CAR-T therapies than White patients. This is a step in the right direction for therapy access when current knowledge shows that Hispanic or Latino patients are less likely to be enrolled in clinical trials and have a higher proportion of patients who live in zip codes with low socioeconomic status and education levels^40^.

Our study revealed that some patients considered eligible for CAR-T therapy did not have a documented discussion about the treatment based on our LLM outputs. Specifically, we found that patients who identified as Other Pacific Islander, Black or African American, and Asian had higher rates of eligibility without a corresponding discussion within the UCSF medical system. These findings suggest that certain patients, particularly those from minority and underserved populations, may be missing out on critical, life-saving CAR-T treatments. However, the relatively small sample size in our analysis represents only the tip of the iceberg, providing a limited view of patients with Multiple Myeloma and highlighting the need for further investigation across other clinical conditions with defined treatment eligibility guidelines.

There are several limitations in our study. ISS staging was unavailable for some patients due to missing beta-2 microglobulin. Therefore, the statistical power of our data was constrained due to missing data on patients’ ISS stage. Due to the inability to extract cytogenetic data, we could not use the updated Revised International Staging System (RISS) staging. Additionally, we could only estimate socioeconomic status at the neighborhood level using ADI, which may have led to ADI not being a significant predictor of CAR-T therapy access. Therefore, we acknowledge that our understanding of socioeconomic status and other social history that can contribute to disparities in access remains incomplete. Our identification of CAR-T eligibility was also limited due to the number of UCSF patients who did not have a clinical note associated with their MM diagnosis and treatments. Furthermore, our study was limited by our understanding of patient enrollment in clinical trials. A future direction of this study would be to incorporate Large Language Models to identify and extract key information from clinical notes concerning SDOH, socioeconomic status, and clinical trial eligibility.

In conclusion, our analysis highlights the association of race, and disease severity on disparities in CAR-T therapy access. Our study shows growing associations of Race on CAR-T treatments regardless of the inverse relationship of worse survival rates in population groups with lower rates of CAR-T administration. However, race is only one way to evaluate disparate treatment for patients and other subgroups based on access or trust in the healthcare system. Furthermore, our study shows that disparities in CAR-T therapy are much more nuanced than therapies associated with other clinical diseases. Because of this nuance, we believe that utilizing individual-level SDOH data could help us dissect disparities at a granular level. Additionally, a deeper analysis of UCSF clinical notes could provide insights into overlooked patients who may benefit from advanced therapies. Differences in access to CAR-T therapies can further disparate the outcomes of MM survival rates in minority population groups. The recognition of disparities within the UC-Health system regarding MM extends to healthcare systems across the state and country. Addressing inequities in CAR-T therapy access is essential to meeting the healthcare needs of MM patients from diverse backgrounds. As treatment protocols evolve and new technologies emerge, it is important to continue investigating disparities in MM care. Ongoing research will help advocate for the equitable allocation of innovative therapies and help increase the survival rates of underserved populations living with Multiple Myeloma.

## AUTHOR CONTRIBUTIONS

Conceptualization, J.D., T.Z., I.Y.C., and A.J.B.; writing—original draft preparation, J.D.; writing—review and editing, J.D., A.K., T.Z., I.Y.C., and A.J.B., Area Deprivation Index data, A.P. All authors have read and agreed to the published version of the manuscript.

## Data Availability

All data produced in the present work are contained in the manuscript

